# 7-Tesla Functional Cardiovascular MR Using Vectorcardiographic Triggering – overcoming the magnetohydrodynamic effect

**DOI:** 10.1101/2021.05.10.21256949

**Authors:** Christian Hamilton-Craig, Daniel Stäeb, Aiman Al Najjar, Kieran O’Brien, William Crawford, Sabine Fletcher, Markus Barth, Graham Galloway

## Abstract

**Objective:** Ultra-high-field (B_0_ ≥ 7 Tesla) cardiovascular magnetic resonance (CMR) offers increased resolution. However, ECG gating is impacted by the magneto-hydrodynamic (MHD) effect distorting the ECG trace. We explored the technical feasibility of a 7T MR scanner using ECG trigger learning algorithm to quantitatively assess cardiac volumes and vascular flow.

**Methods:** 7T scans performed on 10 healthy volunteers on a whole-body research MRI (Siemens Healthcare, Erlangen, Germany) with 8 channel Tx/32 channel Rx cardiac coil (MRI Tools GmbH, Berlin, Germany). Vectorcardiogram ECG was performed using a learning phase outside of the magnetic field, with a trigger algorithm overcoming severe ECG signal distortions.

Vectorcardiograms were quantitatively analyzed for false negative and false positive events. Cine CMR was performed after 3^rd^-order B_0_ shimming using a high-resolution breath-held ECG-retro-gated segmented spoiled gradient echo, and 2D phase contrast flow imaging. Artefacts were assessed using a semi-quantitative scale.

**Results:** 7T CMR scans were acquired in all patients (100%) using the VCG learning method. 3,142 R-waves were quantitatively analyzed, yielding sensitivity 97.6%, specificity 98.7%. Mean image quality score was 0.9, sufficient to quantitate both cardiac volumes, ejection fraction (EF), aortic and pulmonary blood flow. Mean LVEF was 56.4%, RVEF 51.4%.

**Conclusion:** Reliable cardiac ECG triggering is feasible in healthy volunteers at 7T utilizing a state-of-the-art 3-lead trigger device despite signal distortion from the MHD effect. This provides sufficient image quality for quantitative analysis. Other ultra-high-field imaging applications such as human brain functional MRI with physiologic noise correction may benefit from this method of ECG triggering.

**Key points:**

1. Ultra-high field 7 Tesla cardiac MRI is challenging due to the impact of the magneto-hydrodynamic (MHD) effect causing severe distortions in the ECG trace.
2. Using VCG with a learning phase outside the ultra-high field magnet, the R waves can be adequately detected to perform high quality Cardiac MRI scans, overcoming signal distortion from the MHD effect.

## Introduction

Cardiovascular magnetic resonance (CMR) is a increasingly valuable technique for comprehensive morpho-functional evaluation of the left and right ventricles and vascular flow dynamics [1]. Despite their challenges, higher field systems with B_0_ = 3 Tesla are being used in clinical CMR services[2, 3]. Ultra-high-field (B_0_ ≥ 7 Tesla) MR imaging offers further advantages of increased resolution, improved signal-to-noise ratio (SNR) and potentially improved signal contrast and spatial resolution, but at the disadvantage of increased artefacts and difficulties with ECG-gating [3]. Ultra-high field CMR is challenging due to constraints of energy deposition (specific absorption rate, SAR), transmission field non-uniformity, and B0 magnetic field inhomogeneity [4-7]. A major challenge for cardiac imaging at ultra-high field strengths is obtaining reliable ECG gating, which is significantly impacted by the magneto-hydrodynamic (MHD) effect distorting the ECG signal [5, 7].The interaction of a ferromagnetic conductive fluid (blood) within the static magnetic field B_0_ induces a voltage perpendicular to both B_0_ and the direction of blood flow, which is superimposed on the ECG signal, causing substantial derangement of the cutaneous trace [5]. Time-varying magnetic gradient fields also induce voltage perturbations in the ECG leads further distorting the signal [4, 5]. Previous studies of 7T CMR have been often constrained to using pulse oximetry or acoustic triggering [5, 7]. Vectorcardiography (VCG)-based QRS detection algorithms are commonly employed at 1.5 and 3.0 T, which detect the R-wave peak by recognizing the R-wave’s rising amplitude upslope [5, 7]. Other methods of gating include data-driven self-gating and pilot-tone navigation. We explored the technical feasibility of a 7T research MR scanner using a state-of-the-art vector-ECG (VCG) trigger algorithm with a learning phase to create ECG-gated images of left and right ventricles, and aortic and pulmonary vascular flow.

## Materials and Methods

Ultra-high field 7T CMR scans were performed on 10 healthy volunteers using a whole-body research MRI scanner (Siemens Healthcare, Erlangen, Germany) with 8 channel Tx/32 channel Rx cardiac coil (MRI Tools GmbH, Berlin, Germany) under institutional ethics approval (UQ approval 200500050). A detailed discussion of our 7T-CMR acquisition protocol has been described previously, but the present data demonstrate application of this technique for quantitative morphofunctional assessment of both left and right ventricular function and flow quantitation [4]. In brief, we used a breath-held, VCG-triggered retrospectively gated two-dimensional spoiled gradient echo FLASH sequence performed after 3^rd^-order B_0_ shimming with the following parameters: FOV = 360 × 290 mm, matrix = 352×264, thickness = 6.0 mm, TE = 3.1, TR = 63 ms, flip angle = 35 ^°^, parallel MRI (GRAPPA), acceleration factor: 2, reference lines: 24; phases 20. This allowed cine imaging with in-plane isotropic spatial resolution of 1.0 mm and a slice thickness of 4 mm. Full ventricular coverage was performed with sequential short axis slices from apex to base. Steady state free precession (SSFP) imaging was not possible due to specific absorption rate concerns (see below). Two-dimensional phase contrast flow images were acquired in the ascending aorta and proximal pulmonary artery positioned at the level of the sino-tubular junctions.

Vectorcardiogram (VCG) based triggering was performed using a 3-lead wireless ECG trigger device (Siemens Healthcare GmbH, Erlangen, Germany), in conjunction with a matched filter based VCG trigger algorithm [9]. To improve the synchronization performance, the VCG trigger algorithm was calibrated outside of the magnet bore where the MHD effect is negligible [7, 8]. The learning phase of the algorithm was conducted over a period of at least 30 R-R intervals with the subjects lying on the patient table. **Figure 1** shows ECG signals obtained both outside and inside the magnet bore, demonstrating how the trace is substantially altered by the MHD effect causing strong signal distortion. The ST and T-waves are particularly affected, which can lead to an incorrect detection of the QRS complex and mis-triggering of the MR image acquisition. A pulse sensor (Siemens Healthcare GmbH, Erlangen, Germany) was attached to the subjects’ index finger as a backup trigger device.

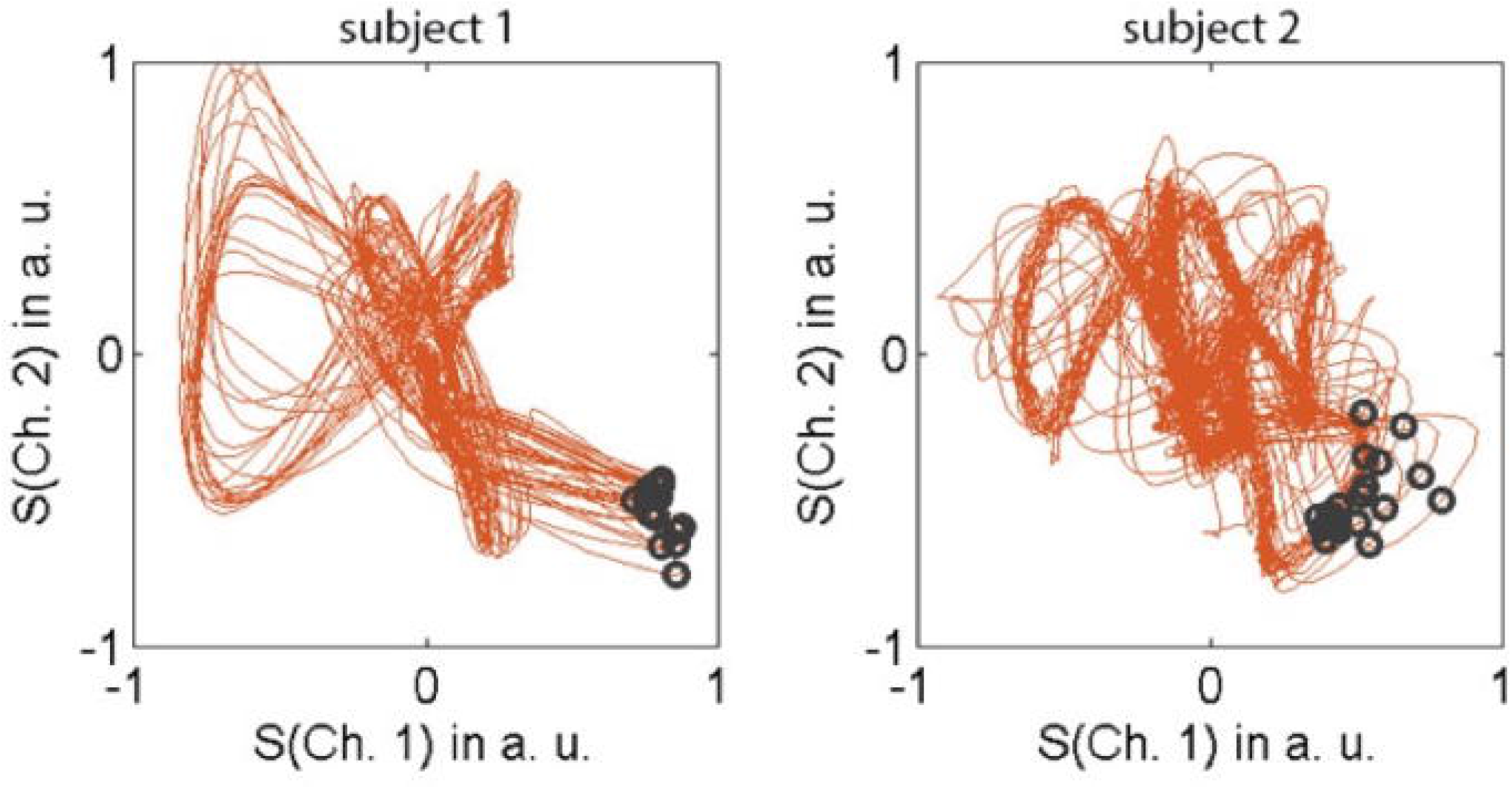

To formally assess the performance of the triggering algorithm, ECG trigger signals and trigger events were recorded. Vectorcardiograms were generated from these data and analyzed quantitatively, **Figure 1**. To obtain a quantitative estimate, false negative (unidentified R-wave) and false positive (triggered by an event that is not an R-wave) trigger events were identified manually in the ECG recordings. From each subject, a representative continuous section of the ECG signal containing up to 500 trigger events was included in the evaluation. From these results, sensitivity and specificity were calculated as follows:

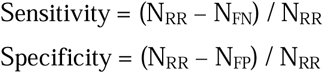

with N_RR_, N_FN_ and N_FP_ denoting the number of RR-intervals, false negatives and false positives, respectively. A representative section of the recorded ECG signal time curves is shown in Figure 1.

Volumetric analysis of cine images and flow quantitation from phase images was performed by a European Association of Cardiovascular Imaging level-3 certified expert reader using cmr42 software (v5.9.4, Circle CVi, Calgary Canada). Rounded ventricular endocardial contours were manually drawn, with the trabeculae and papillary muscles included in the blood pool. [9, 10] Phase contrast flow images were analyzed without background correction. The presence of image artefacts was assessed and graded using a semi-quantitative rating scale from 0 (no artefact) to 3 (severe artefact precluding quantitative analysis).

## Results

VCG-gated 7T CMR imaging was successfully performed in 100% of cases using the learning phase outside of the magnetic field. This resulted in a sufficiently reliable and accurate trigger for CMR acquisition, despite the severe ECG signal distortions from the 7T field (*figure 2*). Quantitative vectorcardiogram analysis of 3142 R-waves was undertaken, yielding 76 false negative (Sensitivity = 97.58%) and 41 false positive (Specificity: 98.70%) events.

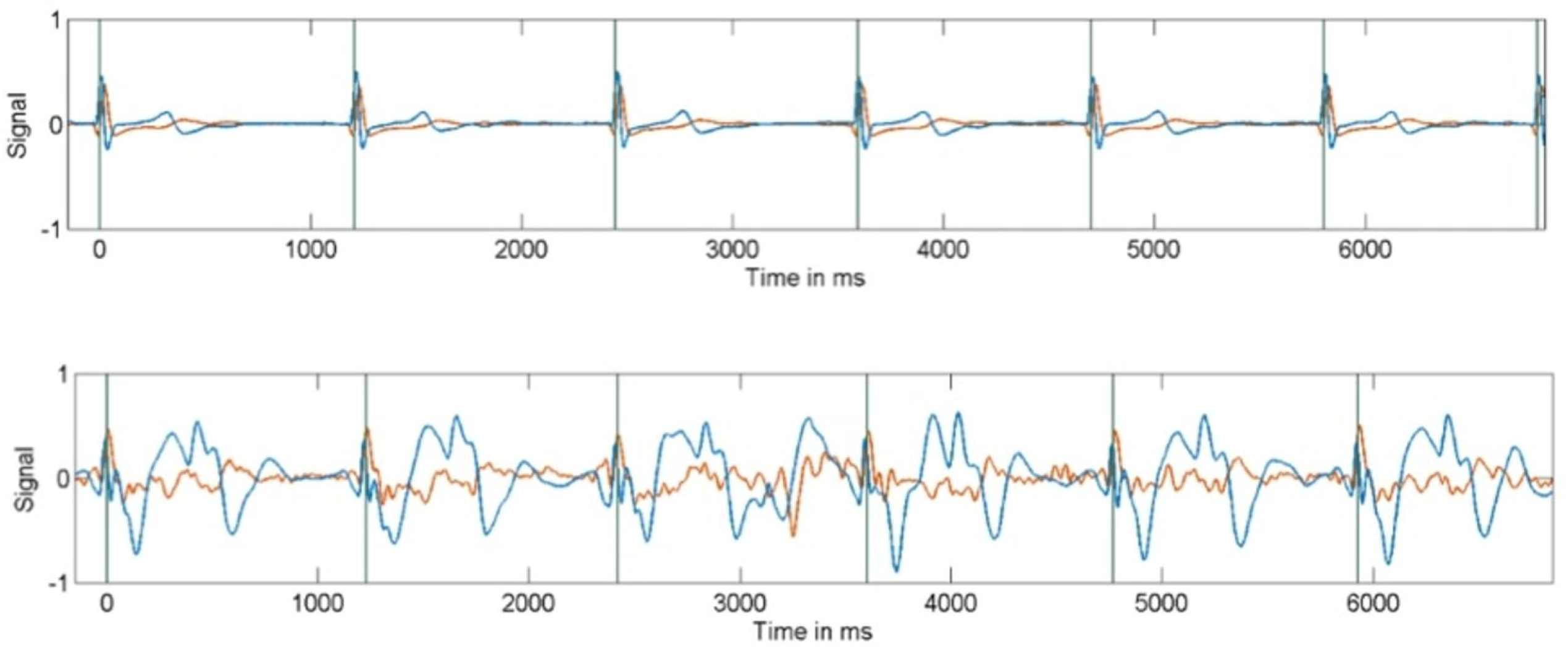

The reconstructed cine CMR images were free of visible trigger-related artefacts, and image quality was sufficient to quantitate both left and right ventricular volumes, ejection fraction, aortic and pulmonary blood flow and regurgitant fractions in all volunteers (*table 1, figures 3-5*). One volunteer had difficulty with breath-holding and a ventricular ectopy, causing mild gating artefacts, which however did not affect quantitative analysis. In 2 other cases, image quality was slightly impaired by signal voids caused by focal RF field non-uniformities **(*figure 2)***, but no case had level 3 artefacts precluding image analysis and volumetric quantitation. Mean image quality score was 0.9 (*table 1)*, indicating very good image quality overall. Mean left ventricular ejection fraction was 56.4%, and mean right ventricular ejection fraction was 51.4% **(*table 1)***.

**Table 1:** Quantitative volumetric analysis of 7T CMR data sets for left and right ventricular volumes, aortic and pulmonary flows

| S<br>u<br>b<br>j<br>e<br>c<br>t | LVE<br>DV<br>ml | LVESV<br>ml | SV ml | LVEF<br>% | Mass g | RV-EDV<br>ml | RV-ESV<br>ml | RV-SV ml | RVEF<br>% | AO-SV<br>ml | PA-SV<br>ml | IQ<br>score |
| --- | --- | --- | --- | --- | --- | --- | --- | --- | --- | --- | --- | --- |
| 1 | 146 | 72 | 74 | 51 | 123 | 167 | 85 | 82 | 49 | 117 | 133 | 1 |
| 2 | 133 | 58 | 75 | 57 | 94 | 136 | 77 | 60 | 44 | 102 | 90 | 0 |
| 3 | 120 | 60 | 59 | 60 | 124 | 134 | 71 | 63 | 47 | 50 | 56 | 1 |
| 4 | 194 | 94 | 100 | 51 | 153 | 205 | 88 | 117 | 57 | 117 | 122 | 1 |
| 5 | 172 | 77 | 96 | 56 | 128 | 195 | 87 | 108 | 55 | 105 | 116 | 1 |
| 6 | 168 | 74 | 94 | 56 | 98 | 148 | 48 | 90 | 61 | 106 | 116 | 0 |
| 7 | 106 | 45 | 61 | 57 | 94 | 139 | 74 | 65 | 47 | 62 | 71 | 2 |
| 8 | 203 | 81 | 122 | 60 | 144 | 200 | 113 | 87 | 44 | 124 | 112 | 1 |
| 9 | 142 | 61 | 81 | 57 | 93 | 157 | 75 | 82 | 52 | 90 | 92 | 0 |
| 10 | 181 | 75 | 106 | 59 | 143 | 183 | 77 | 106 | 58 | 106 | 103 | 1 |
| *m*<br>*e*<br>*a*<br>*n* | *156.5* | *69.7* | *86.8* | *56.4* | *119.4* | *166.4* | *79.5* | *86* | *51.4* | *97.9* | *101.1* | *0.9* |

LVEDV = left ventricular end diastolic volume, LVESV = left ventricular end systolic volume, LVEF = left ventricular ejection fraction, RVEDV = right ventricular end diastolic volume, RVESV = right ventricular end systolic volume, RVEF = right ventricular ejection fraction, AoSV = aortic stroke volume, PASv = pulmonary stroke volume, IQ = image quality artefact score where 0 = no artefact, 3 = severe artefact.

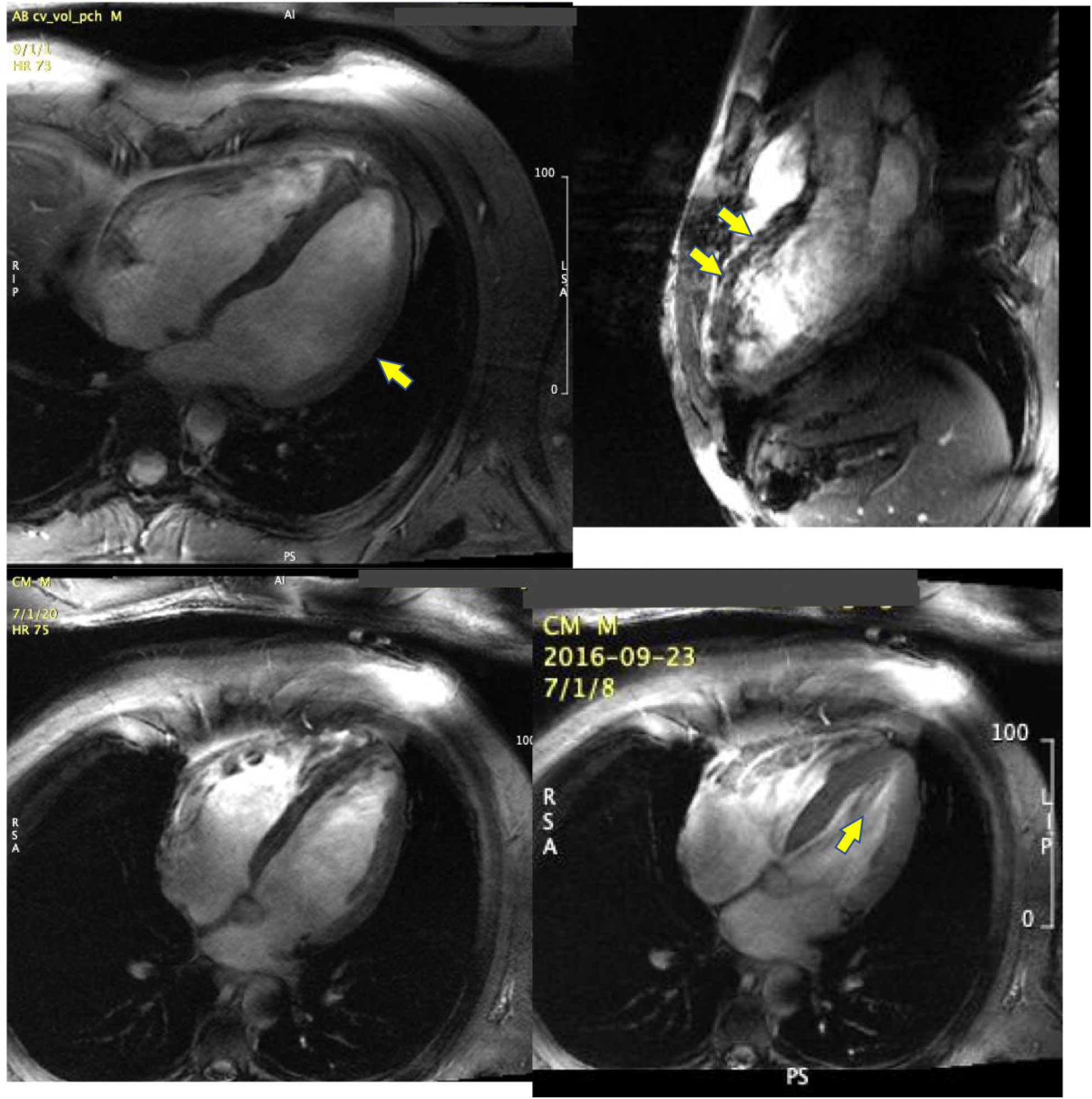

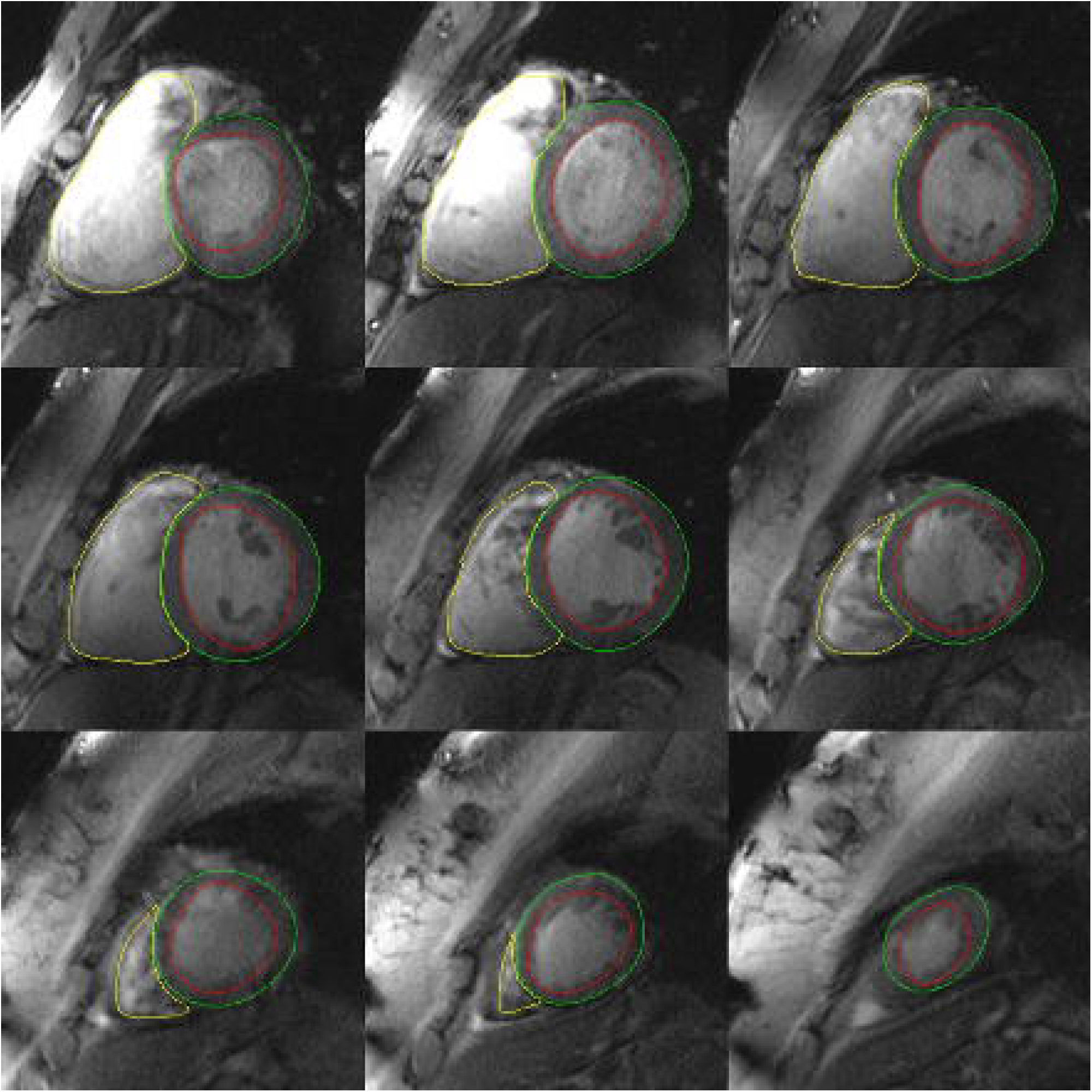

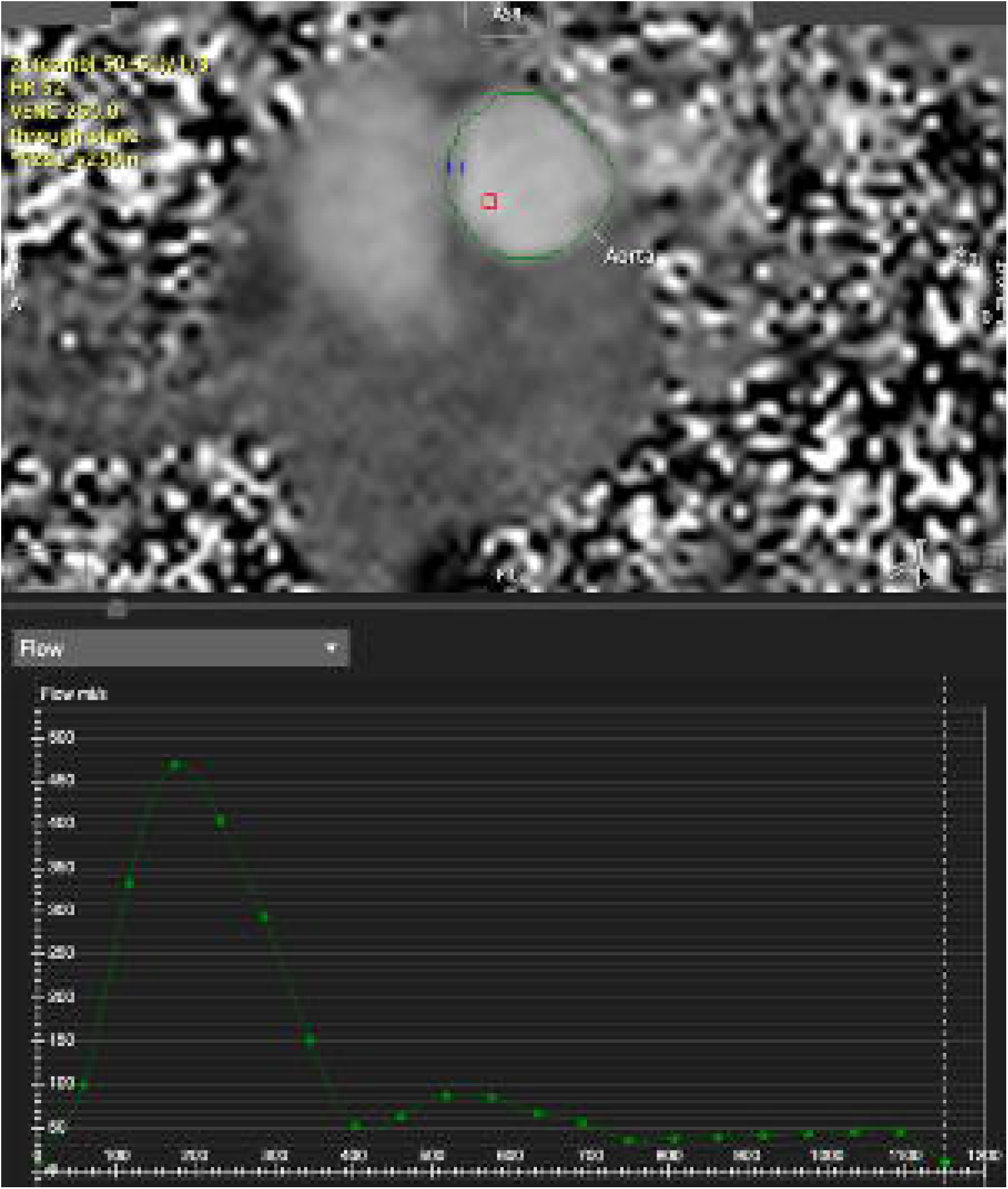

There was excellent correlation between Aortic and Pulmonary stroke volumes, correlation coefficient R = 0.92. There was good correlation between left ventricular stroke volume and aortic stroke volume, R = 0.78, with a bias of -11ml lower left ventricular stroke volume on Bland-Altman analysis, P=0.03.

## Discussion

Vectorcardiogram-triggered imaging, employing an out-of-magnet learning phase, overcame the magneto-hydrodynamic effect on ECG waveforms, allowing acquisition of high quality ECG-gated CMR at 7 Tesla with effective triggering for retrospectively segmented k-space CMR acquisitions.

Electrocardiographic signals are affected by the MHD effect due to the interaction between the blood, a ferromagnetic conductive fluid, and the surrounding magnetic field [5-8, 10]. This interaction causes an electric field distortion which is superimposed on the heart’s intrinsic depolarization, thereby corrupting the signal transferred to cutaneous ECG electrodes. Importantly, the MHD effect is proportional to field strength (B0) and is substantially accentuated at ultra-high field. In addition, the MHD effect is augmented during cardiac systole at the time of maximal aortic blood flow and hence predominantly affects the QRS and T waves. This can cause inaccuracies in detecting the QRS complex and may result in impaired cardiac synchronised imaging. Including a sufficiently long VCG learning phase outside of the magnet bore, where the MHD effect is negligible enabled this problem to be overcome [9, 10], resulted in effective R-wave recognition and successful cardiac synchronised cine imaging at 7T.

We found that VCG-gated 7T CMR was feasible in healthy volunteers, and provided clinically acceptable image quality enabling quantitation of left and right ventricular volumes and systolic function, and aorto-pulmonary vascular flow stroke volumes. Minimal artefacts due to the coil inhomogeneity, off-resonance, motion or a combination of these effects were observed; however, these did not interfere with interpretation and volumetric quantitation (figure 2). The coil inhomogeneity made the right ventricle noticeably more difficult to contour on 7T images than on standard 1.5T and 3T images, with brighter signal anteriorly towards the surface coil and variable signal reduction in posterior myocardial wall (*figure 2, arrow*). In addition, the use of spoiled gradient echo FLASH imaging, rather than steady state free precession (SSFP) which is known to have improved tissue-blood contrast, made ventricular contouring more challenging than standard 1.5/3.0T images. SSFP imaging at 7T has been previously reported by the Oxford group using acoustic gating [6], however the transmit-receive cardiac coil used in our experiments did not allow for SSFP imaging due to exceeding SAR limitations. Parallel transmission coil technology may help to surmount this challenge to allow SSFP imaging at 7T [2].

The small negative bias of lower left ventricular stroke volume compared to the aortic stroke volume calculated from phase contrast imaging can be explained by use of gradient echo FLASH imaging combined with the analysis technique of compacted myocardial contours in these healthy volunteers without valvular dysfunction. The reduced tissue-blood contrast of FLASH imaging compared to SSFP requires “smooth” endocardial contours which ignore trabeculations and papillary muscles [11], thus measuring a larger end-systolic volume and lower stroke volume. This accounts for the small negative bias of stroke volume measured by ventricular contours as compared to phase contrast imaging.

Spatial resolution was improved at 7 Tesla compared to standard CMR imaging. VCG-triggering allowed cardiac synchronised ultra-high field imaging at a slice thickness of 4 mm with an isotropic in-plane resolution of 1.0 mm, in comparison to a slice thickness of 6 to 8 mm and in-plane voxel resolution of 1.2 to 2.0 mm which is common at lower field strengths [12]. Increased resolution may offer advantages in terms of the imaging of thin structures, such as the right ventricular free wall or subvalvular apparatus. SSFP imaging is the clinical standard for CMR imaging and would bring improvements in image quality and tissue-blood borders for volumetric quantitation. However, as discussed above, the use of SSFP was limited by SAR constraints arising from the current coil technology and the need for increased RF power to achieve the same flip angle.

The improved resolution, and reduction in eddy currents at high field strength with gradient shielding, also allowed for very crisp and high quality 2D flow imaging; this has the potential to improve the quantitation of valvular lesions such as aortic and pulmonary stroke volumes and assessment of regurgitation, for which CMR is emerging as the reference standard with improved reproducibility over echocardiographic Doppler-based assessments [13-15]. Gradient echo blood-tissue contrast is improved at 7T compared to standard field strengths, as seen in Figures 2-3. In addition, the increased signal and accurate ECG gating may also allow 4-Dimensional flow (4Dflow) at 7Tesla.

## Limitations

B_1_+ shimming was not conducted, in order to facilitate clinically acceptable examination times for cardiac chamber quantification. The use of B_1_+ shimming, which would be available through parallel transmission techniques, could be beneficial; however, B1+ shimming has implications for signal absorption rate (SAR) management and safety, and was beyond the scope of this experiment.

## Conclusion

Reliable cardiac VCG triggering is feasible in healthy volunteers at ultra-high field utilizing a state-of-the-art 3-lead trigger device with out-of-magnet learning phase, overcoming signal distortion from the MHD effect. This provided sufficient image quality for quantitative analysis. Other ultra-high-field imaging applications such 4D flow and human brain functional MRI with physiologic noise correction may benefit from this method of ECG triggering.

## Data Availability

Data are available upon reasonable request

## Abbreviations

(CMR): Cardiovascular magnetic resonance
(MRI): Magnetic resonance imaging
(SNR): Signal-to-noise ratio
(MHD): Magneto-hydrodynamic
(ECG): Electrocardiogam
(VCG): Vectorcardiography
(GRAPPA): GeneRalized Autocalibrating Partial Parallel Acquisition
(SSFP): Steady state free precession
(RF): Radiofrequency

## Funding

This work was supported by *UQ Academic Title Holder Research Fund and CAESIE: Connecting Australian European Science & Innovation Excellence Priming Grant*.

## Acknowledgement

The authors acknowledge the facilities and the scientific and technical assistance of the National Imaging Facility at the Centre for Advanced Imaging, University of Queensland, Ms Wendy Strugnell FSMRT, the electrophysiology scientists of The Prince Charles Hospital, and funding from the UQ Academic Title Holders grant.

## Data availability

Additional data are available from the Lead/Corresponding author upon reasonable request

## Notes

### Competing Interest Statement

The authors have declared no competing interest.

### Author Declarations

University of Queensland Ethics Review Board UQ approval 200500050

